# An equilibrium solution to the elective waiting list problem

**DOI:** 10.64898/2025.12.29.25343140

**Authors:** Richard M Wood, Sebastian S Fox

**Affiliations:** UK National Health Service (BNSSG Integrated Care Board); University of Bath School of Management; UK Department of Health and Social Care

**Keywords:** Waiting List, Elective Care, Equilibrium Solution, Steady State, Linear Programming

## Abstract

In many countries, demand exceeds supply for elective (non-emergency) hospital treatment, such as hip replacements and cataract removals. The consequence of this is the formation of a waiting list, to which patients join on referral from the family doctor and leave with treatment or ‘renege’ for other reasons (deconditioning, seeking private healthcare, etc). Adequate performance is commonly incentivised through the imposition of targets on waiting times. In the first study to do so, we develop an equilibrium solution to this problem. The specific problem statement is defined based on consultation with senior healthcare managers and policy advisers. After specifying the parameters and dynamical properties, we manipulate the problem into a linear programming formulation. Generally, we find that, based on the amount of reneging deemed tolerable, a continuum of solutions is possible. On application to England’s National Health Service, we quantify the range of solutions in terms of two key metrics: by how much does the waiting list need to reduce and by how much does treatment capacity need to increase. All data and code are publicly available at an online repository. In addition to the results presented here, the solution has been incorporated within a freely available online tool which is in use across several public hospitals as well as local, regional and national planning boards.

## 1. Introduction

At a high level, the elective waiting list problem can be characterised as follows. Patients join a waiting list on referral from a general practitioner. They then wait until hospital treatment, at which point they leave the waiting list. A patient may also leave the waiting list prior to treatment, which is known as ‘reneging’.

Waiting lists are an important area for applied research, given that many healthcare systems worldwide have struggled to keep pace with rising elective demands [Tsui & Fong, 2018; Looi et al, 2023; Hajizadeh & Jalili, 2025]. For instance, in England’s National Health Service (NHS), the elective waiting list has almost doubled in size in the three years from May 2020 to September 2023, and has remained at approximately 7m in the two years since [NHS England, 2025]. Long waits lead to harm for patients [Martinez et al, 2019; Gibbs et al, 2024], additional cost for healthcare providers [James et al, 2024], and reduced economic activity [Williamson & Patel, 2023].

Modelling can improve the clinical, operational and strategic management of waiting lists. Forward projections on an ‘expected’ or ‘do nothing’ basis can allow for more accurate estimates of waiting time to be communicated to patients or their doctors in appropriately managing the condition until treatment. Hypothetical ‘do something’ scenarios, involving perturbations from the ‘expected’ elective demand and capacity profiles, can allow for a higher quality evaluation of interventions – for instance, determining whether a considered initiative is worth the estimated waiting time reduction given its cost. Or managers may use models in an optimisation sense, in order to identify the conditions required for some target on waiting list size or waiting times to be met.

In this paper, our focus is on optimisation: we seek to determine the properties of an elective waiting list which has satisfactory waiting times and is dynamically stable. Specifically, we aim to satisfy the following four objectives:

1. The waiting list must be in equilibrium (or ‘steady state’), such that the characteristics of the waiting list are not fluctuating over time. Ultimately, this means that in a given month, the number of referrals balances the total number of treatments and reneges, and so the waiting list is neither increasing nor decreasing.
2. A set proportion of those on the waiting list must have waiting times within some given threshold. Appreciating that not all treatments can be performed within a fixed period, many healthcare systems mandate that a given percentage must be treated within some target amount of time [Council of Australian Governments, 2011; Rahal et al, 2017]. For instance, in England’s NHS, it is required that 92% of patients on the waiting list have been waiting no longer than 18 weeks [Department of Health and Social Care, 2012].
3. A given proportion of those joining the queue will renege and not be treated. As we will see, it is possible to achieve solutions for the first two objectives with varying amounts of reneging. Reneging, the chances of which generally increase with waiting time, is an interesting phenomenon – it is undesirable from a patient’s perspective, as it could represent having to pay for private healthcare or having become inoperable, but may be advantageous from a healthcare provider’s perspective, as it provides a mechanism to limit waiting list growth without additional expense [Martinez et al, 2019; García-Corchero & Jiménez-Rubio, 2022].
4. The allocation of treatment to those of different waiting durations must be as similar as possible to some given profile. This profile could be based on recent experience and the purpose of seeking similarity is to ensure no radical departure from clinical necessities or behavioural realities. For instance, a 20% allocation of treatments to those in the first month of waiting may be essential to cover more urgent priorities, and so lower allocations may compromise clinical safety. On the other hand, it may be realistically implausible to treat everyone within, say, three months due to the need for pre-surgery weight loss or patients wishing to postpone any major surgery beyond events like weddings or holidays [McLaughlin et al, 2022].

These four objectives have been arrived at through a series of conversations between the authors and healthcare planners and managers in England’s NHS as well as with senior NHS executives and policy analysts within the UK Government. Through these conversations, held through 2024 and 2025, much of the interest had focused on requirements for restoring the aforementioned 92% 18-week target, which had not been achieved at a national level since November 2015 [NHS England, 2025]. Specifically sought were estimations of the required waiting list size to meet this target and the treatment capacity required to maintain it going forward – both at the total national level and for individual regions and clinical specialties. While the application of this study is England’s NHS, the solution developed here is described and specified in general terms and is applicable to any country or healthcare system provided the model remains a suitable approximation of the underlying waiting list dynamics. It should be clarified that this study concerns a strategic view of the elective waiting list problem, in modelling the high-level demand and capacity requirements, and not the bespoke operational workings of a particular waiting list, where the specific consultation, diagnostic and treatment pathways may be considered.

For these more operational-leaning studies, the current literature is relatively developed, including examples for cardiovascular [Catsis et al, 2023], cataracts [Demir et al, 2018] and orthopaedic [Bowers, 2010; Boyle & Mackay, 2022] elective pathways. Most of these studies use Discrete Event Simulation (DES), given its inherent ability to capture variation in the different processes and queues manifesting along the considered pathways. Where larger pathways are studied, the complexity of calibrating the many statistical distributions required for a stochastic DES model can become prohibitive, and necessitate a switch in methodology to deterministic System Dynamics (SD) models. SD models can typically account for a greater range of variables and flows, as well as feedback loops, but can still be used to measure queues and identify bottlenecks – e.g., the only one of the four abovementioned studies to use an SD model concludes “the intervention that would most improve patient flow [along cardiovascular pathways] would be to increase consultant outpatient appointments” [Catsis et al, 2023].

For the more strategic case considered here, the literature is less mature. There are some examples using DES [Comas et al, 2008; Howlett & Wood, 2022], although most efforts appear to recognise the lesser importance of detailed stochastic models when considering higher-level strategy and policy implications, and instead opt for approaches which simply regard the waiting list as some ‘stock’ or ‘compartment’ to which new referrals flow in and treatments flow out [Vissers et al, 2001]. Non-treatment departures (i.e., reneging) from the compartment are also captured [Shah et al, 2024], with some studies modelling this phenomenon by a fixed rate applied to the compartment’s size over time [Hill, 2004; Wood, 2022]. The solutions to these problems (which are essentially basic versions of an SD model) are either through differential-equations if formulated in continuous time or difference-equations if in discrete time. Recently, the single-compartment model has been extended to multiple compartments, where referrals enter the first compartment, representing those in the first month of waiting, and, if not treated or renege within that month, progress, at the turn of the next month, to the second compartment representing those in the second month of waiting, and so on [Wood & Worthington, 2025]. The model, set up in discrete time and solved through a series of iteratively-applied difference-equations, is calibrated using publicly-available NHS data and used to project forward the national waiting list size and shape under various hypothetical time-heterogeneous scenarios for future demand and treatment capacity rates.

This same multi-compartment framework is used in approaching the four modelling objectives of this study, albeit through leveraging a novel mathematical solution which is possible given the simplifying condition of equilibrium (time-homogeneity). It should be noted that some initial results under an early version of this approach have already been published as a short ‘research letter’ (which generated substantial media coverage, given the finding that the national waiting list needed to halve to restore the 92% 18-week target, as pledged by the UK Government [UK Government, 2024]), although this involved a technically inferior solution which appreciated only the first two modelling objectives and included only headline results with limited discussion [Wood et al, 2025]. This current paper explores the full four-objective problem, using a more sophisticated linear programming formulation, and delves much deeper into the model dynamics and resulting insights.

The remainder of this paper is structured as follows. In Section 2, we describe the problem statement in terms of the compartmental framework and detail its mathematical solution. Section 3 contains the results on application of the model to England’s NHS, where we focus on the aforementioned matters of required waiting list size and ongoing capacity requirements. Finally, we discuss strengths, limitations and further research opportunities in Section 4.

## 2. Methods

Overall, the approach is to determine the properties required to satisfy the abovementioned four objectives. Mathematical formulae are developed for application to a waiting list at any chosen level of granularity, e.g., at a national total level or for a particular specialty at a particular hospital – provided that the relevant input data exists. The three main inputs required are: the referral rate, specifying the number of referrals that join the waiting list in a given unit of time; the renege parameters, specifying the probability of reneging based on elapsed waiting time; and a given treatment allocation profile, specifying the allocation of treatment to those waiting different durations. Additionally required are specification of the waiting time threshold (e.g., 18 weeks) and quantile (e.g., 92%) and the target reneging rate (e.g., 15%).

The outputted waiting list properties will consist of the required size of the waiting list and its shape, in terms of the numbers waiting different durations, and the required amount of treatment capacity and its allocation profile, in terms of how treatments are allocated to those waiting different durations. In comparing to the current state of any studied waiting list, the following metrics are considered as overall indicators of the shorter and longer term effort required: the required one-off reduction in waiting list size, subtracting the required equilibrium waiting list size from the current waiting list size; and the required recurrent expansion in treatment capacity, subtracting the current treatment capacity from the required equilibrium treatment capacity.

### 2.1 Compartmental framework

The waiting list is modelled by the one-way progression of patients through a series of compartments representing increasing waiting time (Figure 1). Let *λ* > 0 represent the number of newly referred patients that join the first compartment in each time-step. We assume a time-step of one month henceforth. At each compartment and in each month, waiting patients are exposed to the chances of treatment or reneging, both resulting in departure from the waiting list. The remaining patients flow into the next compartment at the following month, i.e., all compartments ‘empty’ into the next with each passing month. Following treatments and reneging, let *n*_*m*_ represent the number of referrals in the *m*-th compartment representing those in their *m*-th month of waiting. Let *T*_*m*_ ≥ 0 represent the number of treatments of those in their *m*-th month of waiting. Reneging occurs as a proportion 0 ≤ *r*_*m*_ ≤ 1 of the inflowing patients to each compartment at each month, so the numbers reneging in their *m*-th month of waiting is *r*_*m*_*n*_*m*−1_. This is a simplified version of the compartmental modelling framework developed by Wood & Worthington [2025] (the simplification is to time-homogenise the parameters given the equilibrium results sought here).

**Figure 1.**
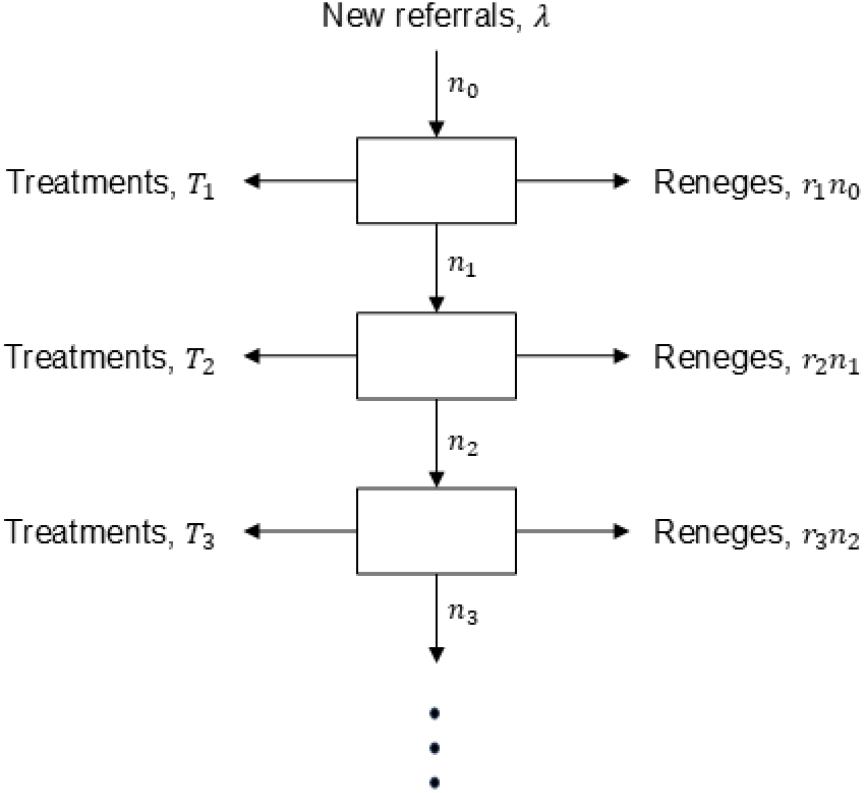
Compartmental modelling framework used to represent the waiting list problem.

Of the *λ* = *n*_0_ new referrals entering the first compartment in a given month, *T*_1_ are treated and *r*_1_*λ* renege such that *n*_1_ = *λ* − *r*_1_*λ* − *T*_1_ = *λ*(1 − *r*_1_) − *T*_1_ remain and pass through to the second compartment at the following month. Of these, *T*_2_ are treated and *r*_1_*n*_1_ renege such that *n*_2_ = *n*_1_(1 − *r*_2_) − *T*_2_ remain and pass through to the third compartment at the following month. And so on.

Expanding the *n*_*m*_ for the first four terms:

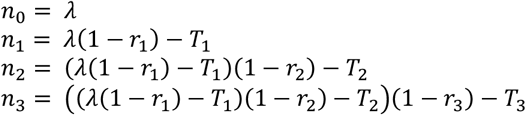

Separating and collecting the *λ* and *T* terms yields:

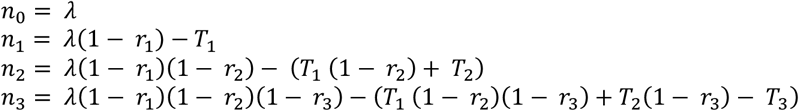

This generalises to:

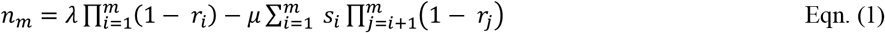

for *m* = {0,1,2, … }, noting that 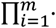 for *m* < 1 is the empty product (equal to one) and 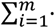 for *m* < 1 is the empty sum (equal to zero).

### 2.2 Objectives

The four aforementioned objectives (Section 1.1) can now be expressed in terms of the compartmental modelling framework.

#### Objective one: equilibrium

To ensure equilibrium, the waiting list must not be increasing nor decreasing and so the number of referrals per month must be equal to the total number of treatments and reneges:

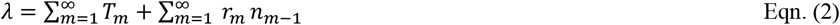

#### Objective two: waiting times

To ensure that a proportion of those currently waiting are waiting under a certain duration:

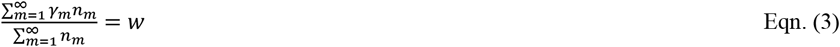

where 0 < *w* < 1 is the proportion and the vector 0 ≤ *γ*_*m*_ ≤ 1 indicates the threshold duration. E.g., *γ*_*m*_ = {1,1,1,0,0,0,0, … } would correspond to a requirement whereby *w* of those waiting must be waiting no longer than three months (in such case, 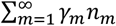 is the number waiting up to three months while 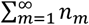 is the total number waiting). Eqn. (3) can be simplified to:

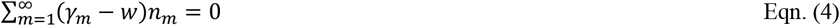

#### Objective three: reneging

To ensure that total reneging equals some set level:

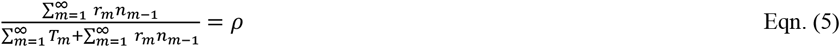

where *ρ* > 0 is the given proportion of all waiting list departures that are reneges. Eqn. (5) can be simplified to:

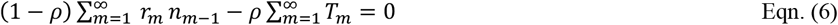

#### Objective four: treatment allocation profile

To minimise the differences between the treatment allocation profile and a reference one (presumably based on recent historical data, for reasons previously given):

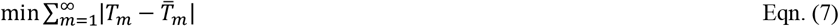

where 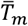 is the reference treatment allocation profile (scaled such that 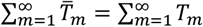). Note that the treatment allocation profile also, of course, provides the distribution of total time to treatment (this is not to be confused with the instantaneous distribution of waiting time for those currently waiting when observed at a particular moment in time, which is given by the *n*_*m*_).

### 2.3 Linear programming problem

We want to solve Eqns. (2), (4), (6) and (7) for the unknown treatment allocation vector *T*. We do so by coercing the problem to a Linear Programming (LP) formulation consisting of three equality constraints covering the first three objectives and an optimisation part covering the fourth objective.

#### 2.3.1 Treatment of Equations (2) and (6)

Observe that in Eqns. (2) and (6) there is the term *r*_*m*_*n*_*m*_ − 1, which, on substitution of Eqn. (1), can be written:

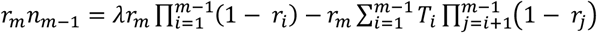

and so:

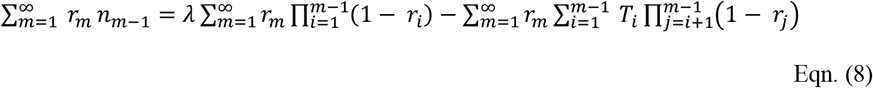

Consider only the *T* part:

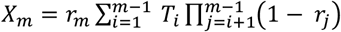

Letting *R*_*j*_ = (1 − *r*_*j*_), the first five terms are:

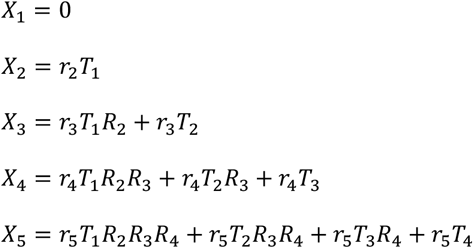

Collecting the *T*_*m*_ terms, we can see:

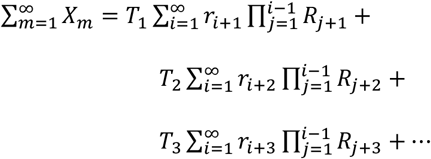

Hence:

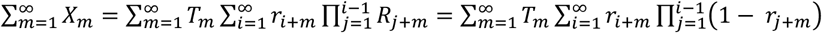

and therefore, returning to Eqn. (8):

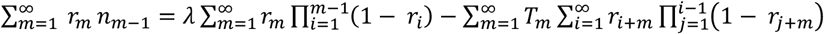

Note that the terms 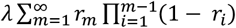 and 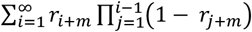, for a given *m*, are constants since *λ* and *r* are known.

For ease, we set 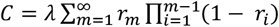 and 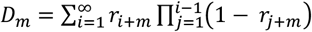, hence:

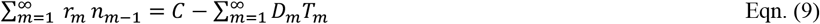

With Eqn. (9) we can now write Eqn. (2) as:

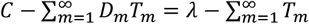

and so:

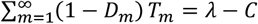

Setting *E*_*m*_ = 1 − *D*_*m*_ and *F* = *λ* − *C* we have:

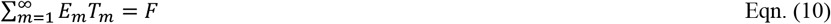

With Eqn. (9) we can now write Eqn. (6) as:

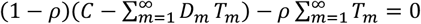

and so:

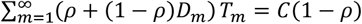

Setting *G*_*m*_ = *ρ* + (1 − *ρ*)*D*_*m*_ and *H* = *C*(1 − *ρ*) we have:

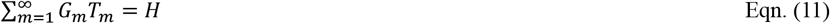

#### 2.3.1 Treatment of Equation (4)

Observe that in Eqn. (4) there is the term (*γ*_*m*_ − *w*)*n*_*m*_ which, on substitution of Eqn. (1), can be written:

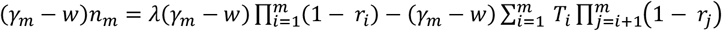

and so:

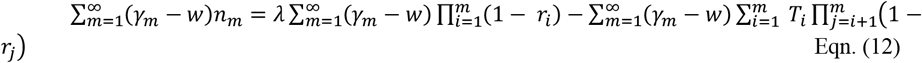

Consider only the *T* part:

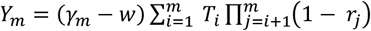

Letting *R*_*j*_ = (1 − *r*_*j*_), the first four terms are:

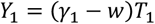

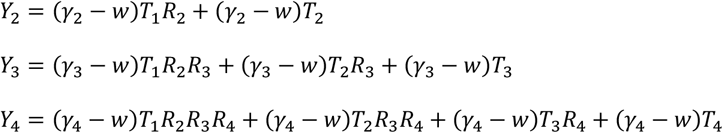

Collecting the *T*_*m*_ terms, we can see:

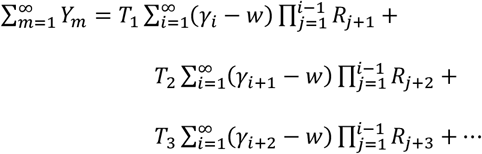

Hence:

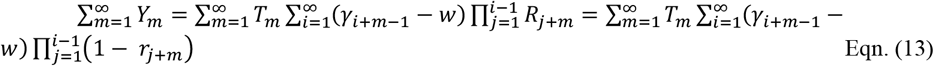

and therefore, returning to Eqn. (12):

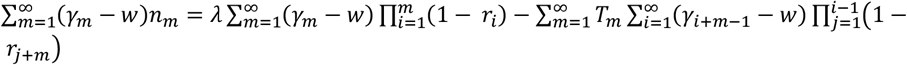

Note that the terms 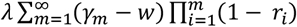 and 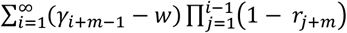, for a given *m*, are constants since *λ, w, r* and *γ* are known.

For ease, we set 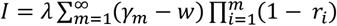 and 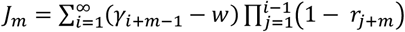, hence:

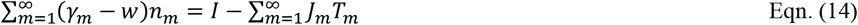

With Eqn. (14) we can now write Eqn. (4) as:

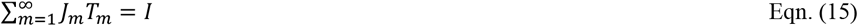

#### 2.3.3 Treatment of Equation (7)

Introduce auxiliary variables *t*_*m*_ ≥ 0 such that 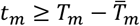 and 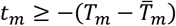. The objective now is:

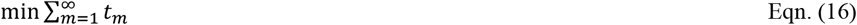

#### 2.3.4 Problem statement

We want to solve the LP problem:

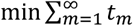

such that:

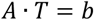

where:

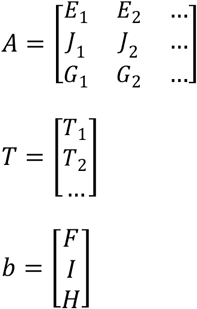

and all unknown variables are non-negative, i.e., all *t*_*m*_ ≥ 0 and *T*_*m*_ ≥ 0.

#### 2.3.5 Problem statement, under finite m

Clearly, the infinite case is impractical: one cannot estimate the *r*_*m*_ for very large *m*, and neither does one need to since the contributions to the formulae will become negligible as – realistically – the vast number of referrals will either be treated or renege before reaching substantial waiting times. For instance, at year-end 2024, only 155 of the 7.5m on England’s NHS waiting list were waiting beyond 24 months [NHS-E, 2025].

Hereon we bound *m* such that *m* = {0,1,2, …, *M*}. Practically, the choice of *M* should reflect the consequences of being too large or too small. As mentioned, setting it too large may be unnecessary and may mean having insufficient data to reliably calibrate the parameters (potentially leading to spurious parameterisations). On the other hand, consider the dynamical implications of imposing a finite upper summation limit on *m*: any referrals remaining on the waiting list beyond that time are ignored – from the perspective of the formulae, they are simply no longer on the waiting list despite having neither been treated nor reneged. Clearly, this is implausible. An appropriate *M* is therefore large enough for the waits of only a negligible number of patients to exceed such a duration while also being small enough to provide sufficient data to achieve reliable parameterisation.

Under finite *m*, bounded above by *M*, we therefore want to solve the LP problem:

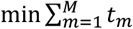

such that:

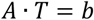

where:

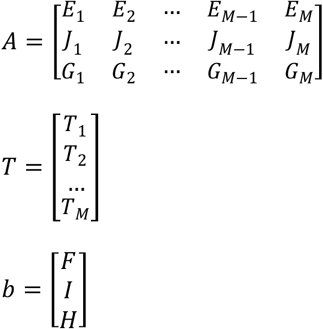

and:

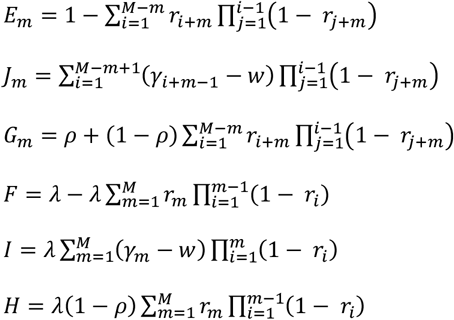

and all unknown variables are non-negative, i.e., all *T*_*m*_ ≥ 0 and *t*_*m*_ ≥ 0.

#### 2.3.6 Solution

Considering the equality constraints *A* · *T* = *b*, there are (presuming *M* > 3) more unknowns than equations, and so the problem is underdetermined and will have either no solution or infinitely many solutions. If the latter, the minimisation objective ensures that a single solution is reached. The solution is obtained through the use of the ‘lp’ function within the ‘lpSolve’ package in the statistical programming language R [Csardi & Berkelaar, 2024]. This function requires the following inputs:

- *direction*: character string giving direction of optimisation: ‘min’ (default) or ‘max’.
- *objective*.*in*: numeric vector of coefficients of objective function.
- *const*.*mat*: matrix of numeric constraint coefficients, one row per constraint, one column per variable.
- *const*.*dir*: vector of character strings giving the direction of the constraint: each value should be one of ‘<‘, ‘<=‘, ‘=‘, ‘==‘, ‘>‘, or ‘>=‘.
- *const*.*rhs*: vector of numeric values for the right-hand sides of the constraints.

We now step through how each of these are set in turn:

- *direction*: set to ‘min’, as we have a minimisation problem.
- *objective*.*in*: set to {{0}^*M*^, {1}^*M*^}, i.e., a 2*M* length vector with *M* zeroes followed by *M* ones, as we want to minimise the 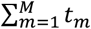.
- *const*.*mat*: set to 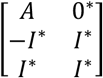, which is a (3 + 2*M*) × 2*M* matrix and where *I*^*^ is the *M* × *M* square identity matrix and 0^*^ is the 3 × *M* zero matrix.
- *const*.*dir*: set to {{′ = ′}^3^, {′ ≥ ′}^2*M*^}, to represent the three equality constraints and 2*M* inequality constraints with the auxiliary variables introduced in Section 2.3.3.
- *const*.*rhs*: set to 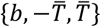, i.e., a 3*M* length vector.

Note that in the ‘lpSolve’ R package, all unknown variables are assumed to be non-negative as default, and so no additional constraints are required to ensure *T*_*m*_ ≥ 0 and *t*_*m*_ ≥ 0.

### 2.4 Example application to England’s NHS

As mentioned earlier, the numbers awaiting elective treatment in England’s NHS have doubled in recent years, with – at the time of writing – just 62% of the current 7m waiting list waiting under 18 weeks [NHS England, 2025]. This is far less than the 92% requirement [Department of Health and Social Care, 2012], which the current Labour government has pledged to restore by 2029 [UK Government, 2024].

Using the model, we ask what size the waiting list must reduce to in order to realise this ambition and what amount of ongoing treatment capacity would be required. As well as investigating this for the total national waiting list, we also consider regional and specialty level requirements. To begin with, we permit various amounts of reneging in order to help understand the model dynamics as well as to reflect the practical uncertainty in what level of reneging is tolerable. For further experiments, we restrict to *ρ* = 0.1, i.e., a 10% renege rate (or the largest amount of reneging up to 10% where a feasible solution can be found). In terms of the other parameters, we set *M* = 12 and, given the 92% 18-week target (where 18 weeks is 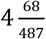 months), *w* = 0.92 and 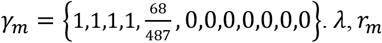 and 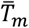 are both estimated from the publicly-available data for financial year 2024/25, i.e., from April 2024 to March 2025 inclusive [NHS England, 2025].

### 2.5 Online repository

The data and code (in R) used to implement the model and generate the results presented here are publicly available at the online repository ‘https://github.com/nhs-bnssg-analytics/waitlist_equilibrium_model’. This repository also includes the full modelled results.

## Results

Table 1 presents some of the key properties of the elective waiting list in England as of the 2024/25 financial year. At a national level, the largest backlog is for Trauma and Orthopaedic (682,032), followed by Ear, Nose and Throat (590,625), Gynaecology (541,711) and Ophthalmology (526,344). Mean times to treatment vary from 2.7 months (Elderly Medicine) to 5.0 months (Neurology) and all estimated rates of reneging are above 15%. Within England, the Midlands has the longest waiting list (1,230,809), followed closely by London (1,229,205) which, at 4.0 months, has the longest mean time to treatment. Specialty-level detail for the regions can be found at the aforementioned online repository.

**Table 1.**
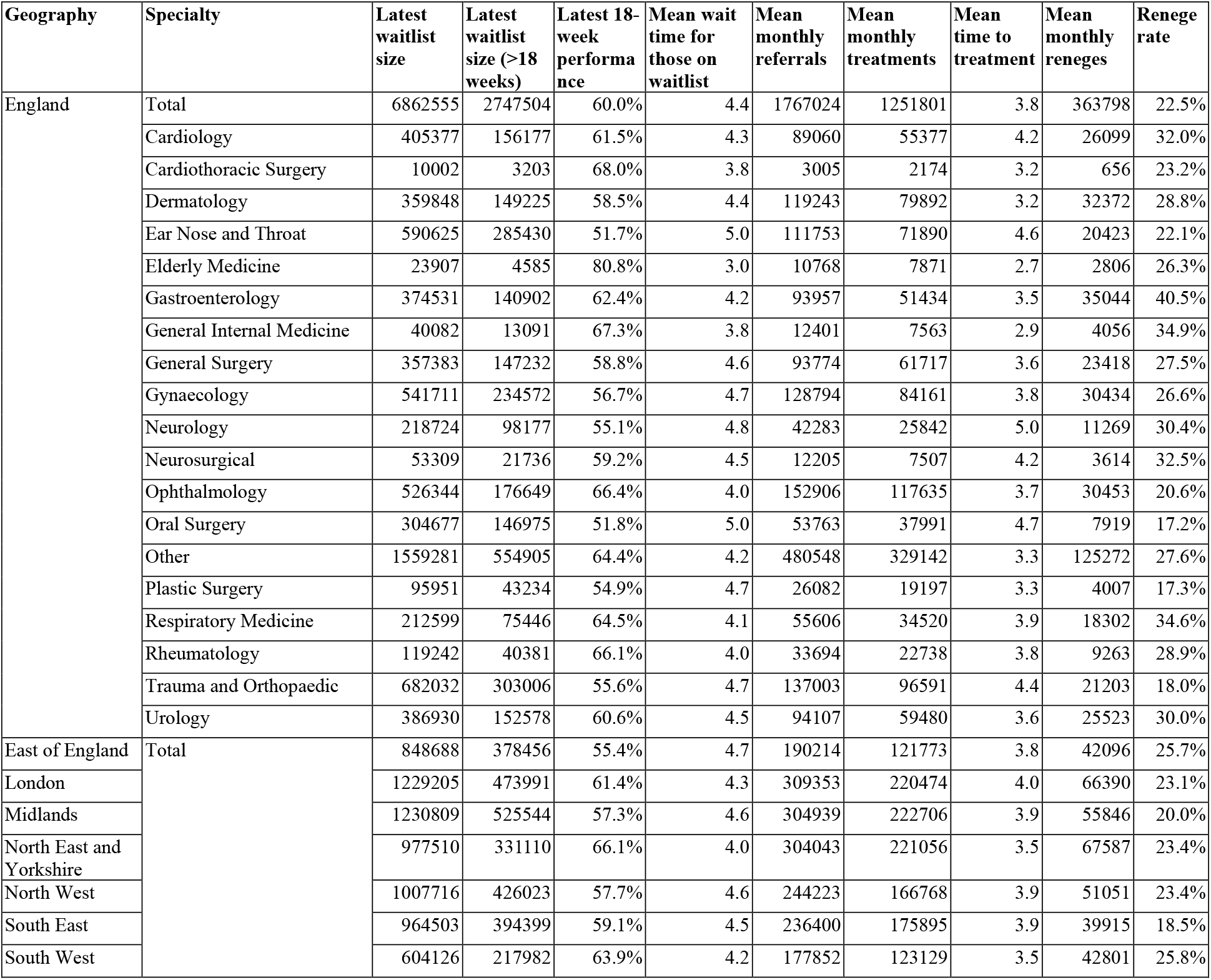
Descriptive summary of the data used for modelling, covering the 2024/25 financial year.

Figure 2 shows the modelled results for the total England waiting list. Specifically, eight results were found corresponding to specified renege rates of 2.5% to 20% by increments of 2.5%. All of these represent solutions to the problem formally described in Section 2.2 (equilibrium; 92% 18-week performance; closest treatment allocation profiles). Larger renege rates were attempted but no feasible solutions were found beyond 20%. This means that, under current reneging behaviour (bottom-left panel), an equilibrium solution achieving the 92% 18-week target cannot be achieved with the current 22.5% renege rate (Table 1), and so – recognising that referrals must balance treatments and reneging – more treatment capacity is required. So, as would be expected, lower renege rates associate with more treatment capacity (top-left panel). We also see that smaller waiting lists correspond to lower levels of reneging, which is achieved through prompter treatments (top-right panel). At higher levels of reneging, it becomes necessary to keep more patients on the waiting list for longer; essentially to provide them with greater opportunity to renege and so to ensure satisfaction of the specified renege rate. This results in a longer waiting list and an increasing mass of required treatment allocation to those in the fifth month of waiting. This required ‘point mass’, combined with lesser treatment allocation to those in earlier months, is strictly necessary to ensure that sufficient numbers both renege and are removed shortly beyond 18 weeks when they no longer contribute to the ‘numerator’ of the 18-week calculation (Eqn. (3)) and so would otherwise depress the value below the 92% target. It should also be noted that, while all solutions achieve 92% performance, the absolute number of waiting patients who are waiting over 18 weeks is, of course, proportionate to the total waiting list size, which increases with the specified renege rate (top-left panel). The mean times to treatment (which correspond to the treatment allocation distributions in the top-right panel) also increase, from 1.5 months (2.5% renege rate) to 5.0 months (20% renege rate), compared to the current 4.4 month mean (Table 1). Tabulated results for all regions and specialties are available at the online repository.

**Figure 2.**
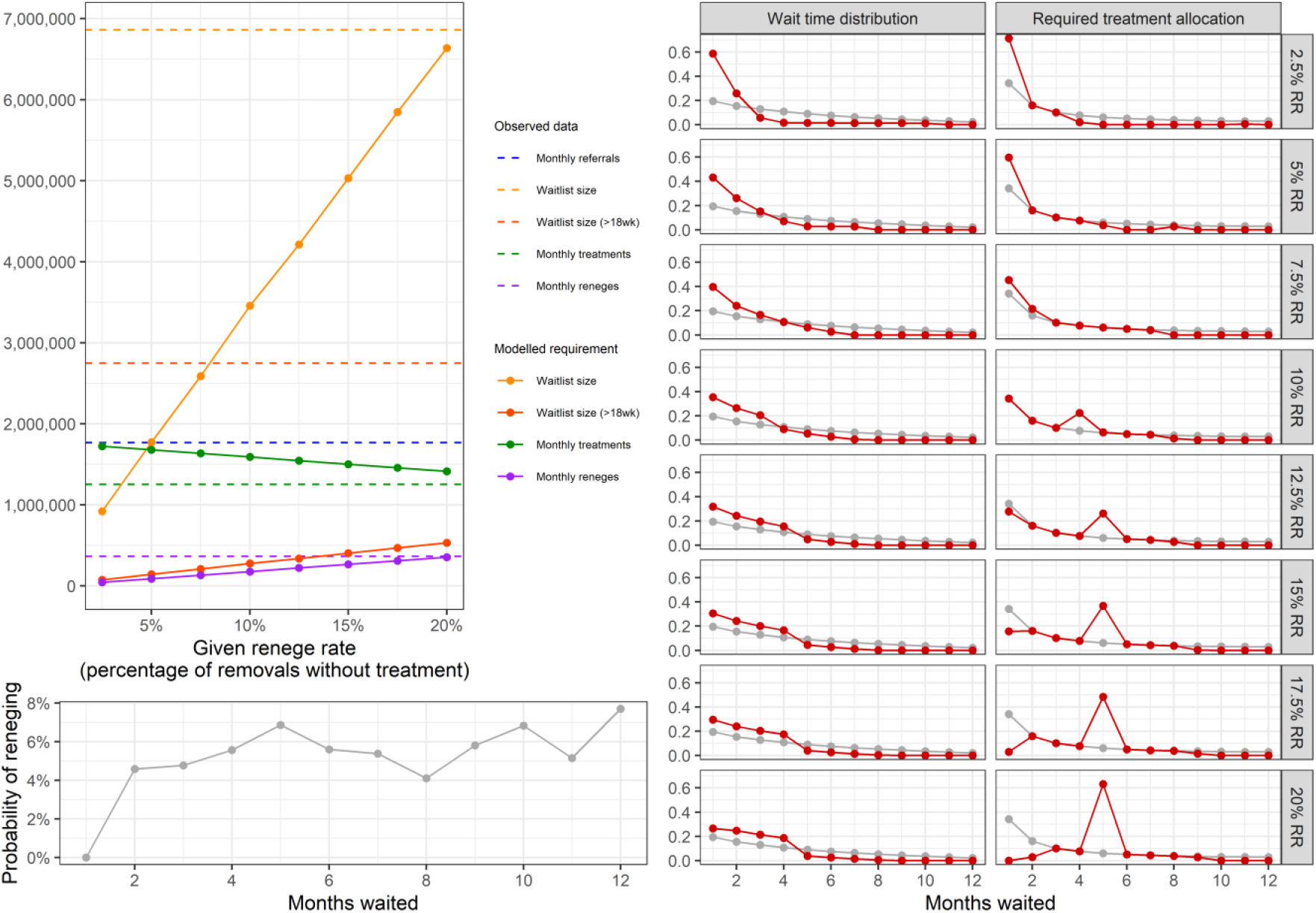
Modelled results for the total waiting list for England to achieve equilibrium and 92% 18-week performance. Where not otherwise indicated, observed data for 2024/25 is shown as dark grey with modelled results as dark red. ‘RR’ is renege rate.

Figure 3 shows, for each region and specialty and for different renege rates (5%, 15% and 25%), the estimated non-recurrent (one-off) waiting list reduction and extra recurrent (ongoing) treatment capacity required to achieve an equilibrium waiting list meeting the 92% 18-week target. Generally, these challenges are lessened at higher rates of reneging, for reasons already explained. It was not possible to find solutions for some problems, e.g., only one region’s Oral Surgery waiting list was feasible at 15% reneging (North East and Yorkshire) and none at 25%. Where multiple solutions are possible across the considered renege rates, observe the differences in the scale of the two challenges of reducing the waiting list and expanding capacity in response to varying renege rates for different region’s waiting lists within given specialties. For instance, the proportionate differences appear similar among regions for Dermatology (the coloured lines have similar angles) whereas General Surgery in the South West requires proportionately greater treatment capacity expansion than waiting list reduction with reducing reneging rates (steeper angle for the pink line) when compared to the Midlands (shallower angle for the dark green line). As well as the angles, note also differences in the length of the lines, e.g., compare North East and Yorkshire versus South West for Neurosurgical. These phenomena are being driven, in part, by differences in the reneging profiles (*r*_*m*_) between the specialties and regions.

**Figure 3.**
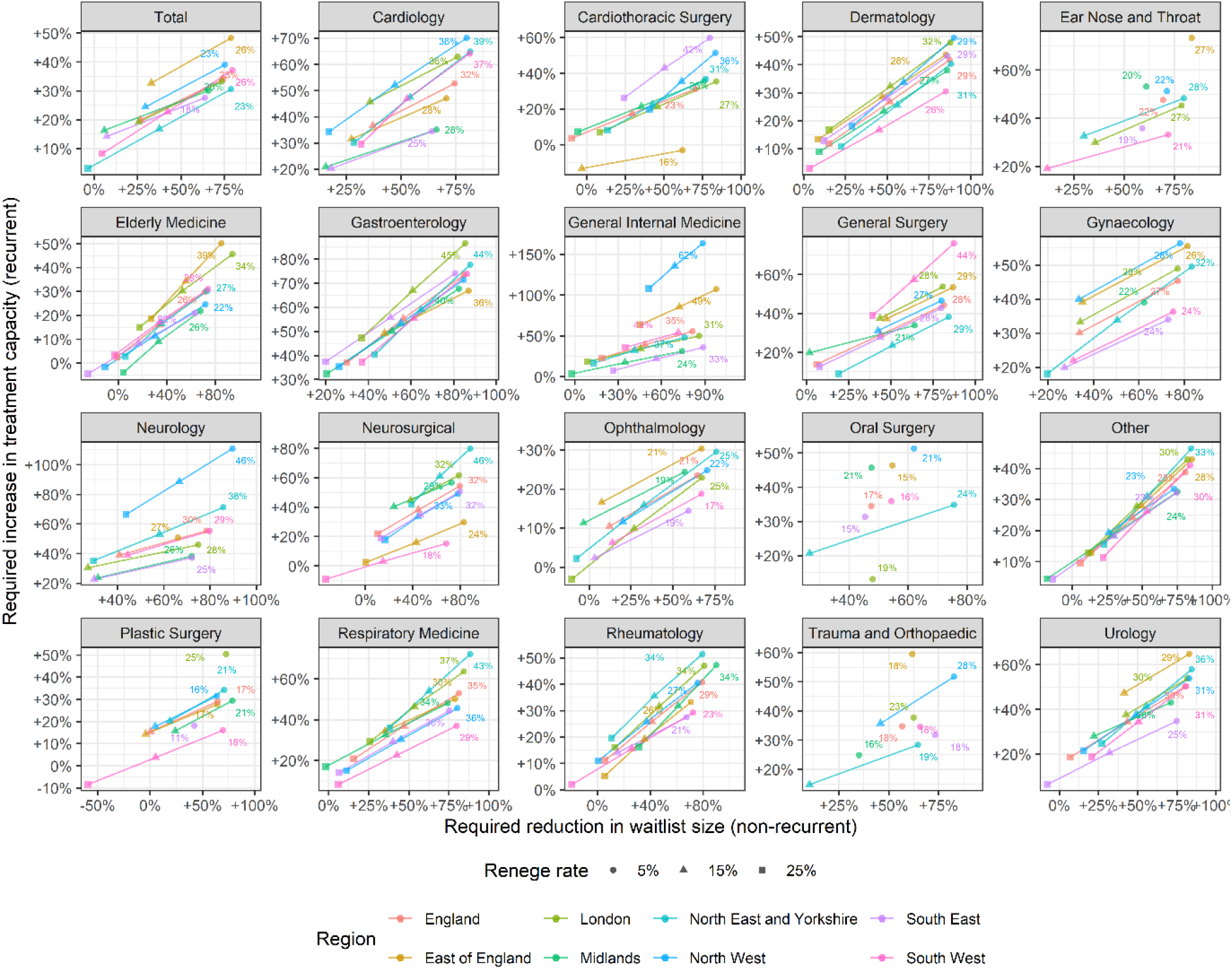
Estimated scale of the challenge required in reducing waiting lists and expanding treatment capacity to achieve equilibrium and 92% 18-week performance. The overlaid numbers included in each panel detail the current reneging rates.

Figure 4 describes the regional distribution of these challenges, under the studied 10% renege rate. This illustrates the unequal balances of the two challenges. For instance, Midlands and North East and Yorkshire require similar treatment capacity expansions (23% and 24% respectively) but the former requires only a 35% waiting list reduction while the latter requires a 58% reduction. Figure 5 provides a specialty-level breakdown, from which it can be seen that Dermatology and Gastroenterology require the largest reductions in waiting list (70% and 71% respectively, at the national level), but while Gastroenterology also requires the largest increase in recurrent treatment capacity (64%), Dermatology requires a more modest 34% increase, which is fairly standard on comparison to the other specialties. Other noteworthy results include recurrent treatment capacity reductions (not increases) for Cardiothoracic Surgery (East of England and South West) and substantial regional variation in required treatment capacity increases for General Internal Medicine (24% for Midlands versus 150% for North West).

**Figure 4.**
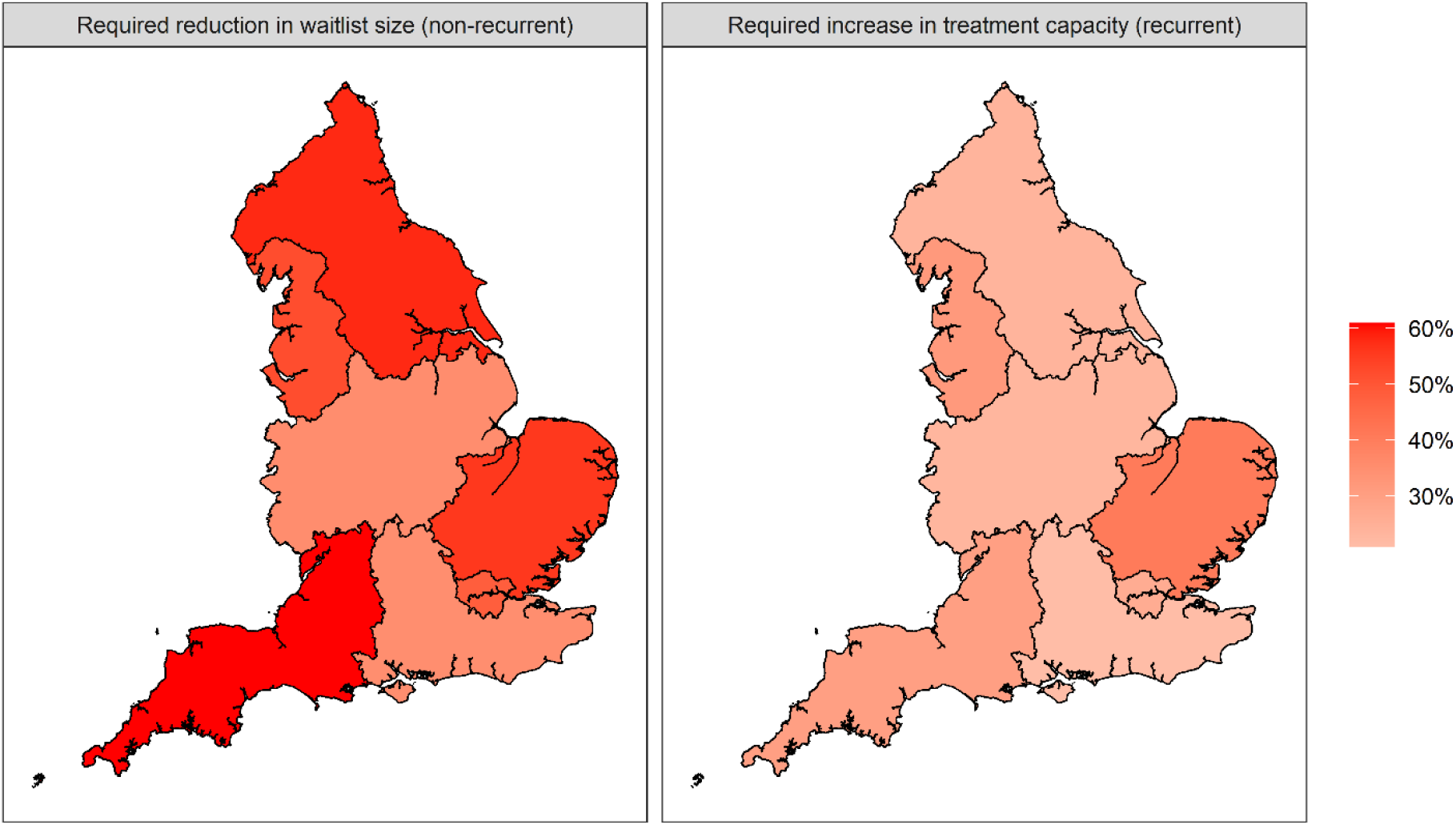
Estimated scale of the challenge required in reducing waiting lists and expanding treatment capacity to achieve equilibrium and 92% 18-week performance for 10% reneging. Results are for the total specialty waiting lists for each region in England.

**Figure 5.**
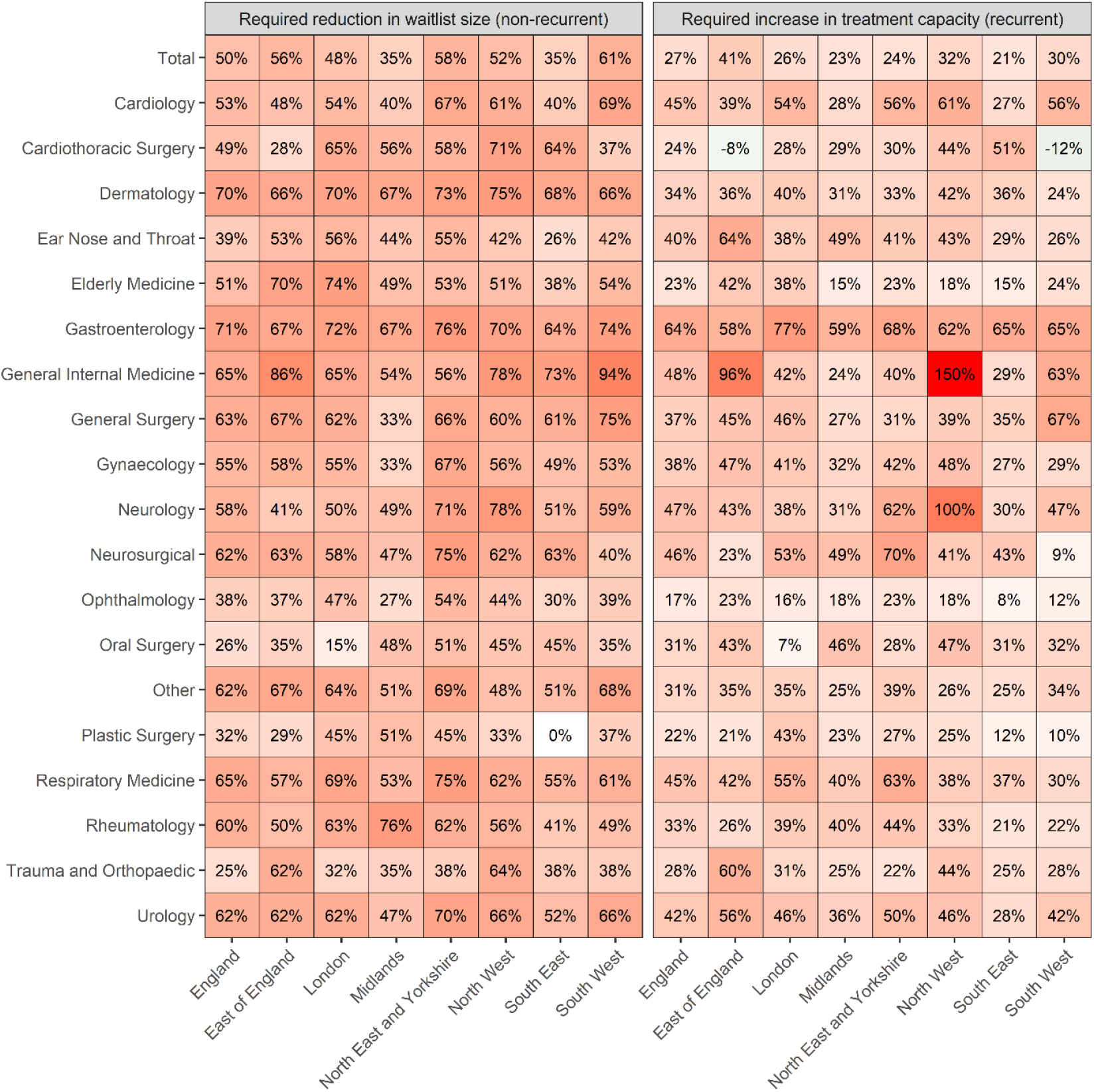
Estimated scale of the challenge required in reducing waiting lists and expanding treatment capacity to achieve equilibrium and 92% 18-week performance for 10% reneging. Results are for the waiting lists at national and regional level and for each specialty.

Figure 6 shows the differences in the modelled versus current proportions of treatment capacity allocated among different waiting durations, under the same 10% reneging rate. The exhibited ‘point mass’ of required treatment capacity to those in the fifth month of waiting and variation in month one required allocation manifest for the same reasons as previously raised for the national example of Figure 2. In line with other reported results, there appears substantial differences between the regions and specialties, with some particularly large and consistent month five spikes for the Ear Nose and Throat, Oral Surgery, and Trauma and Orthopaedic specialties, and spikes for the Midlands (General Surgery) and East of England (Neurology). The practical significance of these variations is discussed later in Section 4. Figure 7 illustrates a key reason for the month five spikes, which relates to the level of reneging experienced in the first four months (approximately 18 weeks) of waiting. Essentially, if fewer patients have reneged during this period, there is more of a need to remove them when they pass 18 weeks and so would otherwise exit the numerator (of Eqn. (3)) and depress performance below the required 92%. This association is visually evident from Figure 7 and supported by linear modelling of these two components (p<0.0001).

**Figure 6.**
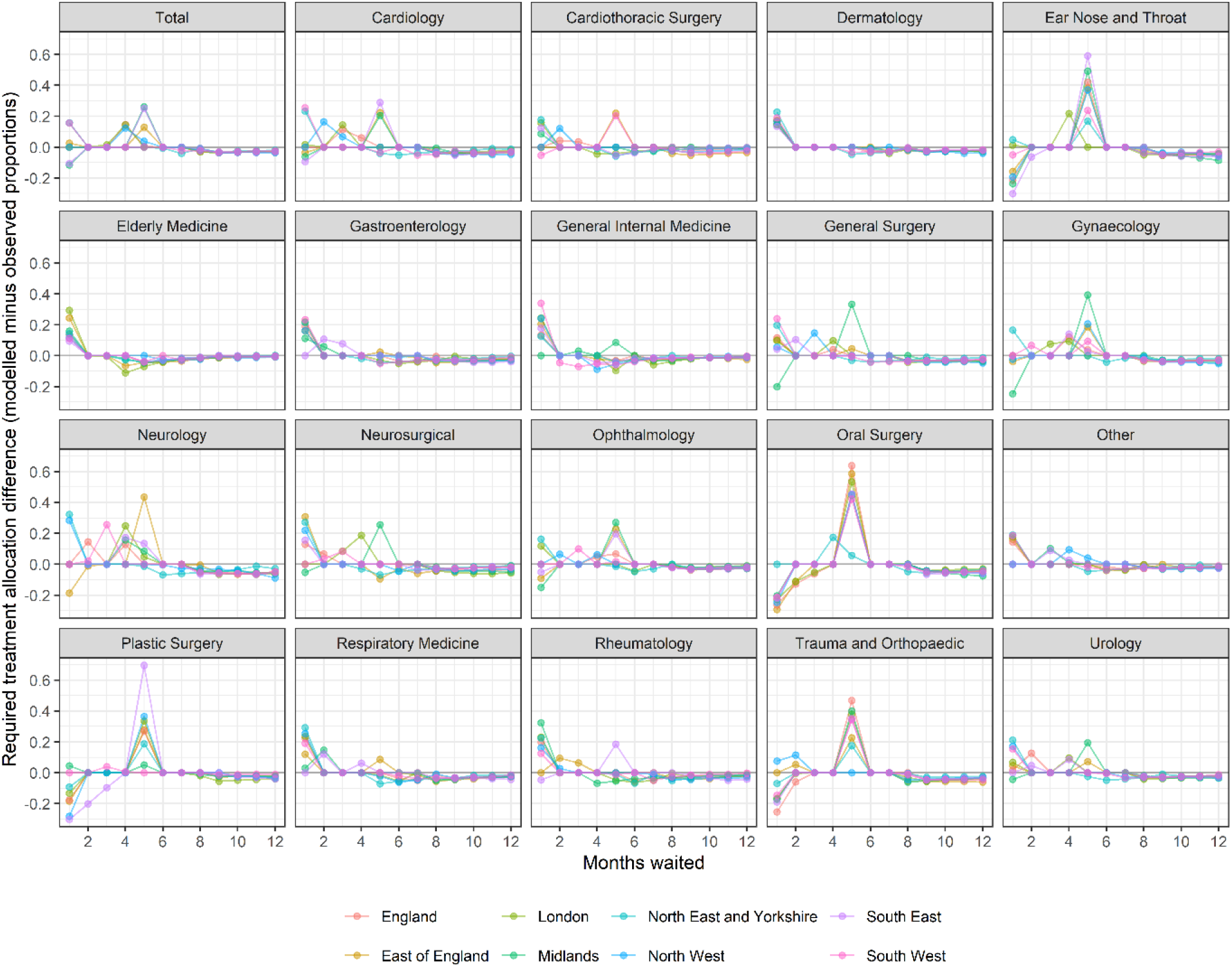
Differences in the modelled versus current observed proportions of treatment capacity allocated among different waiting durations for 10% reneging.

**Figure 7.**
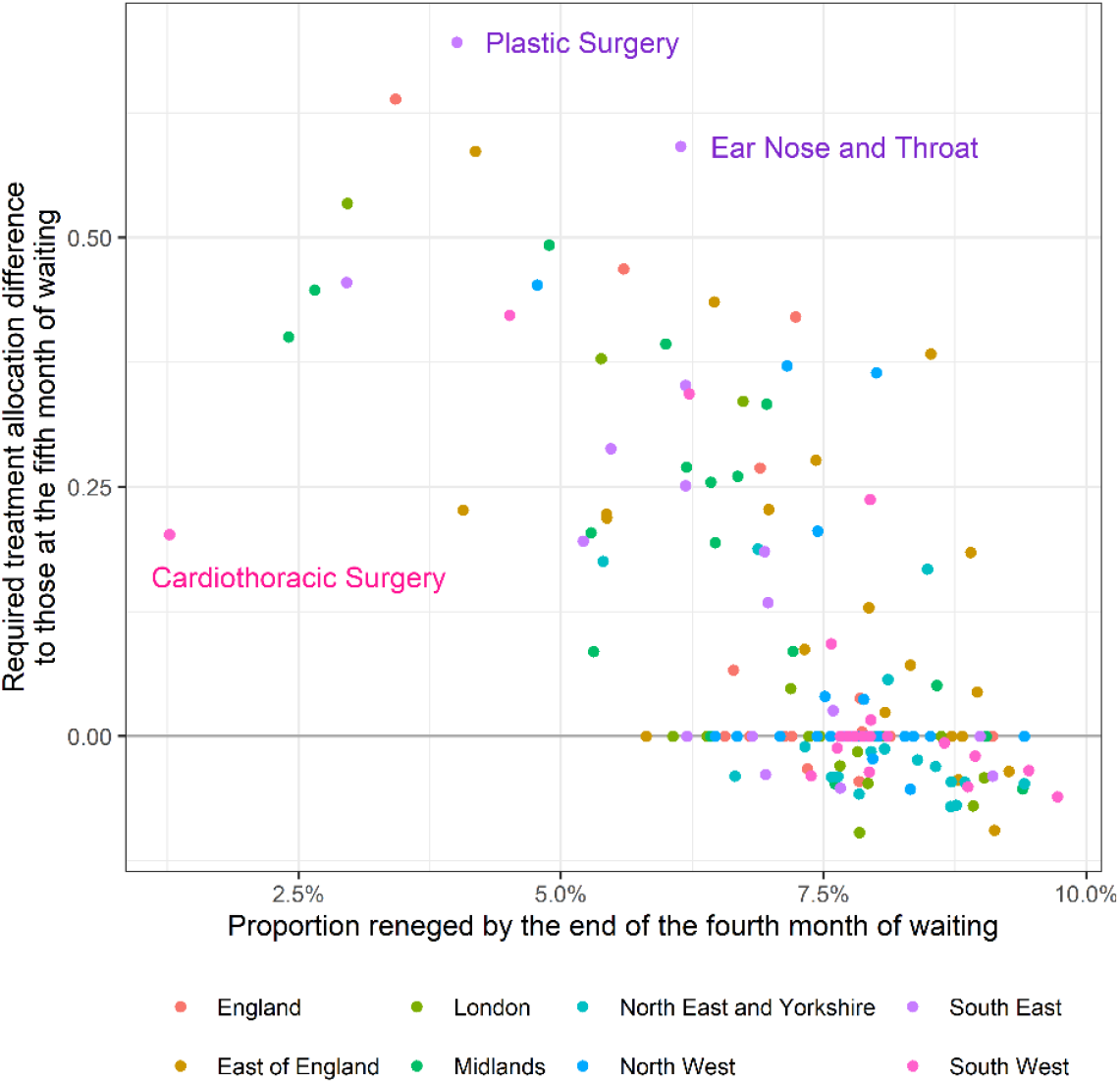
Association between the proportion of referrals that renege by the end of the fourth month of waiting and the difference in modelled versus current treatment allocations to those in the fifth month of waiting.

## 4. Discussion

The aim of this study was to derive an equilibrium solution to the elective waiting list problem, insofar as satisfying the four objectives arrived at through discussion with senior healthcare managers: steady state, some set target on waiting time, a set rate of reneging, and the closest match to some given treatment allocation profile. Using a compartmental modelling framing of the waiting list dynamics (Section 2.1), these objectives (Section 2.2) were solved using a linear programming method (Section 2.3), which was applied – as an example – to the elective waiting list for England’s NHS (Section 2.4). An equilibrium solution to the elective waiting list problem as specified here has not before been studied and so the findings of this work represent a novel contribution to the academic literature.

This study also has important practical implications. As previously remarked, there is a deficit of information to guide healthcare administrators in understanding how a waiting list works and how it can be improved, particularly with regards to strategic aspects of the problem. The concept of an ‘equilibrium’ waiting list provides managers with a clear aiming point in specifying the properties that need to be achieved. This conveniently gives rise to the two key metrics considered here which inform management how much effort is required in the short-term, in reducing the waiting list size, and recurrently, in ensuring sufficient ongoing treatment capacity to maintain performance. In addition to the results contained in this paper, it should be noted that the solution has also been implemented within an interactive planning tool that has been developed by the authors for use in England’s NHS [NHS BNSSG ICB, 2025]. The freely-available tool, which is hosted on a server and accessible over the internet, is known to be in use by several hospitals trusts and integrated planning boards, as well as regional oversight functions (a blog documenting the development of the tool has already been published [NHS-R Community, 2025]).

Practically, this study raises some possible concern regarding the use of a performance target based on the waiting times of those currently waiting, such as the one considered here (at least 92% of currently waiting patients must be waiting no longer than 18 weeks). The ‘spikes’ in required treatment capacity allocation to those in the fifth month of waiting is an indicator of possible trouble. This appears to be required at higher reneging rates, as it becomes necessary to deliberately extend the waiting time of patients to provide more opportunity for them to renege. This has parallels with healthcare providers ‘gaming’ waiting time targets used in emergency departments, with allegations of “hitting the target but missing the point” [Mason et al, 2012; Tenbensel et al, 2019]. This also means longer waiting lists, and, while the 92% target may be met, the target has no recognition of the absolute numbers waiting over 18 weeks, e.g., for the England total waiting list, the absolute numbers waiting over 18 weeks double if 20% reneging can be tolerated over 10% (Figure 2). As previously mentioned, this will have consequences for patient harm [Martinez et al, 2019; Gibbs et al, 2024] and lead to additional cost for healthcare providers [James et al, 2024]. Moreover, it would also lead to longer times to treatment: from a mean wait of 2.9 months (10% reneging) to 5.0 months (20% reneging), for the national waiting list. Again, it is worth reaffirming that under both of these options the 92% target remains met.

In terms of limitations, the utility of an equilibrium solution itself should firstly be appraised. Practically, of course, the waiting list and its constituent parts are never constant – there will be periodic fluctuations due to the number of working days each month, seasonality in available hospital treatment capacity (lower in winter months due to higher non-elective demands), and trends over time relating to demographic change and advancements in treatments. That said, while England’s NHS has seen remarkable variation in recent years, with 18-week performance dropping from 83% to 47% in the first five months of the Covid-19 pandemic, it has remained between 57% and 64% from January 2022 to September 2025 and had been within five percentage points for the six-year period from March 2011.

Another limitation – and where some further research may be required – is in regard of referral priority. In practice, referrals are allocated to one of a number of priority classes based on how urgently the treatment is required. This is difficult to capture within a model ‘explicitly’, since different specialty waiting lists have different priority classes and these can change over time, and – moreover, at least for this study’s application – no such data is publicly reported. Instead, the compartmental modelling framework attempts to capture priority ‘implicitly’ through the treatment allocation profile, i.e., a greater mass in month one may represent the more urgent treatments. However, it may also just be indicative of a waiting list that has not been under pressure and so has been able to more promptly perform treatments. The problem here is not knowing how much of a waiting list is ‘urgent’ and within what timeframe such treatments are required. This is a problem because, while the fourth objective promotes a close match to recent treatment allocation, the closest match (which satisfies the first three objectives) may still be substantially different. Observe the large difference in the proportion of treatment capacity allocated to those in the first month of waiting as reneging is increased from 2.5% to 20% for the national waiting list as considered in Figure 2 – from a clinical perspective, are the solutions for 15% reneging and higher even feasible, on account of performing substantially fewer potentially-urgent month-one treatments? Further work may explore the appropriateness of introducing further constraints to restrict the space of solutions considered feasible.

Another complexity worthy of further research regards the amount of reneging deemed tolerable. Here, we investigate multiple solutions through varying the rate of reneging as per the third objective. Essentially, this is premised on there being no published target reneging rate in England’s NHS and there existing a multitude of reneging rates observed in practice (Table 1). Of course, management does not directly set the amount of reneging but influences it through the treatment allocation profile, and this study reports the degree of correspondence between the two. However, unless and until there is any formally set target on reneging or further work to understand what levels may be tolerable, it will be difficult to provide managers with a single equilibrium solution. This, from a practical perspective, is challenging since, while the findings of this study may have educative value in explaining and quantifying the dynamical intricacies of a waiting list, any inability to provide single figures for the required waiting list reduction or treatment capacity increase will inevitably limit the extent of practical use in planning exercises where straightforward inputs are necessary. Further work is therefore required to consider what levels of reneging may be clinically or politically tolerable and the degree to which such levels depend on treatment specialty.

## Data Availability

This data uses publicly available data hosted here: https://www.england.nhs.uk/statistics/statistical-work-areas/rtt-waiting-times/. The processed data is available here: https://github.com/nhs-bnssg-analytics/waitlist_equilibrium_model.

## Acknowledgements

The authors are grateful to Lucy Morgan and Neil Walton for their contributions to the early thinking on the need for an equilibrium solution to this problem.

